# Re-Irradiation of Recurrent Head and Neck Cancers Using Pulsed Reduced Dose Rate Radiotherapy: An Institutional Series

**DOI:** 10.1101/2024.01.17.24301218

**Authors:** Romy Megahed, Arpan V. Prabhu, Delanie P. Mack, Somayeh Gholami, Santanu Samanta, Mausam Patel, Gary D. Lewis

**Affiliations:** Department of Radiation Oncology, University of Arkansas for Medical Sciences, 4301 W. Markham St., #771, Little Rock, AR 72205-7199

## Abstract

**Purpose/Objective(s):** Pulsed reduced dose rate (PRDR) radiation (RT) is a re-irradiation (Re-RT) technique that potentially overcomes dose/volume constraints in the setting of previous radiation therapy. While the use of this Re-RT technique has been reported for other disease sites, there is minimal data for its use for recurrent or secondary primary squamous cell carcinoma of the head and neck (HNSCC). In this study, we report preliminary data from our institution of a consecutive cohort of patients with HNSCC who received PRDR Re-RT.

**Materials/Methods:** Out of 11 patients who received PRDR Re-RT from August 2020 to January 2023, 9 had analyzable data. Intensity modulated RT was used for treatment delivery using either volume modulated arc therapy or helical tomotherapy. A wait time between 20cGy arc/helical deliveries was used to achieve the effective low dose rate. Data collected included patient demographic information, prior interventions, diagnosis, radiation therapy dose and fractionation, progression free survival, overall survival, and toxicity rates.

**Results:** The median time to PRDR from completion of initial RT was 13 months (range, 6-50 months). All but one patient underwent salvage surgery prior to PRDR. In total, 4 patients received systemic therapy as part of their re-treatment courses. The median follow-up after Re-RT was 7 months. The median OS from PRDR was 7 months (range, 1-32 months). Median PFS was 7 months (range, 1-32 months). One patient (11.1%) had acute grade 3 toxicity, and two patients (22.2%) had late grade 3 toxicities. There were no acute or late grade 4 or 5 toxicities.

**Conclusion:** PRDR for Re-RT is a feasible treatment strategy for patients with recurrent or second primary HNSCC. Initial findings from this retrospective review suggest reasonable survival outcomes and potentially improved toxicity; prospective data is needed to establish the safety and efficacy of this technique.

## Introduction

Despite progress in radiotherapy (RT) delivery, up to 40% of patients with squamous cell carcinoma of the head and neck (HNSCC) will either develop a locoregional recurrence or second primary.^1^ Salvage surgery is a potential curative option for these patients, but many patients are not suitable candidates.^2^ Systemic therapy may improve survival but is not a curative option.^3^ Re-irradiation (Re-RT) is a curative treatment option but is associated with significant risk of toxicity.^4^

There has been interest in advanced radiation techniques including stereotactic body radiotherapy (SBRT) and proton therapy to reduce toxicity and side effects of Re-RT by decreasing the volume of normal tissue irradiated. Pulsed Reduced Dose Rate (PRDR) RT is an alternative Re-RT technique that improves the therapeutic ratio of Re-RT, not by decreasing the amount of normal tissue irradiated, but by changing its biologic impact. PRDR-RT delivers low-dose pulses of RT that can cause hyper-radiosensitivity of tumor cells and increased DNA damage repair of normal tissue cells.^5,6^ Previous research has shown encouraging results for Re-RT using PRDR-RT in multiple disease sites.^7–9^ However, there are minimal reports on the use of PRDR-RT for HNSCC. The purpose of this study was to report preliminary efficacy and toxicity data from a single institution of a consecutive cohort of patients with HNSCC who received Re-RT via PRDR-RT.

## Materials and Methods

A retrospective review of patients who received Re-RT at our institution from August 2020 to January 2023 for HNSCC (recurrent or second primary) using PRDR-RT was conducted. A waiver of informed consent was provided by our institutional review board due to the minimal risk posed to study participants. Patient demographic and clinical treatment information was collected and de-identified. Acute and late toxicity were classified by retrospective chart review and scored using the Common Terminology Criteria for Adverse Events (CTCAE), version 5.0. Pre-existing feeding tube or tracheostomy dependence was not considered toxicity nor was tracheostomy use after laryngectomy. Toxicity events developing after locoregional recurrence were considered disease-related and not included. Progression free survival (PFS) was defined as progressive disease or death. Overall survival (OS) was defined as death from any cause. Survival outcomes were calculated from the last day of PRDR Re-RT.

### PRDR-RT technique

Re-RT was targeted to the location of gross disease (definitively or post-operatively) with an expansion of 7-10mm for clinical target volume (CTV) while also considering the extent of the surgical bed per physician discretion. Elective nodal radiation was not performed. A 3mm planning target volume (PTV) was added. Daily setup included an Aquaplast facemask and computed tomography. For all plans, 95% of the PTV was covered by 100% of prescribed dose. Plan evaluations were based on dose–volume histograms, the equivalent dose at 2Gy, treatment durations, and dose–volumes to normal tissues. Dose to normal tissues was kept as low as reasonably achievable in accordance with consensus guidelines.^10^ Intensity Modulated Radiation (IMRT) was used for treatment delivery using either volume modulated arc therapy (VMAT) or helical tomotherapy. A wait time between 20cGy arc/helical deliveries was used to achieve the effective low dose rate of ∼6-7cGy/min.^11^ For helical tomotherapy, the pitch factor and the gantry period were determined in a way to move the couch slowly through the gantry aperture with a constant speed; the slow movement of the couch between the 20cGy helical passes provided enough time to achieve the effective low dose rate. For VMAT, a timer was used to ensure enough time (3 minutes) between each 20cGy arc.

## Results

### Patient characteristics

In total, 11 patients received HNSCC Re-RT via PRDR-RT. However, 2 patients did not complete treatment for unrelated reasons (transportation issues, mechanical fall resulting in injury) and were not included in the final analysis. Patient characteristics and previous treatment details for the 9 analyzed patients are outlined in Table 1. All patients received prior RT before PRDR Re-RT; median dose of prior standard RT was 70Gy (range, 63-70Gy) given with 2-2.25Gy per fraction. Three patients had prior surgery and/or chemotherapy as part of their initial treatment course. The median time to PRDR Re-RT from completion of initial RT was 13 months (range, 6-50 months).

**Table 1:**
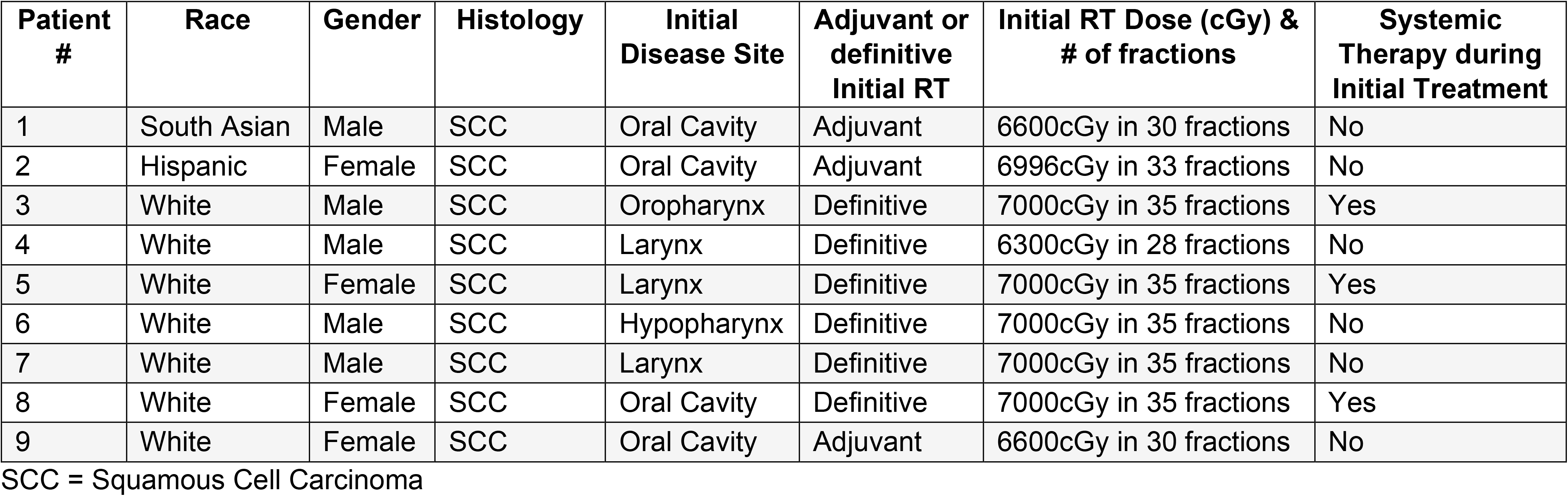
Patient Characteristics & Previous Treatment Details.

### PRDR Re-RT

All patients underwent multidisciplinary discussion at our institution’s head/neck tumor conference prior to PRDR Re-RT. Re-RT treatment details for the 9 analyzed patients are outlined in Table 2. All but one patient underwent salvage surgery prior to PRDR Re-RT; indications for adjuvant PRDR Re-RT are included in Table 2. 60Gy in 30 fractions was prescribed to the patients who underwent salvage surgery and 66Gy in 33 fractions was prescribed to the patient who received definitive PRDR Re-RT. CTV volumes ranged from 19.1 to 234.1cc with three patients have CTV volumes >100cc. In total, 5 patients received systemic therapy as part of their re-treatment courses, either concurrently and/or adjuvantly with respect to PRDR Re-RT.

**Table 2:** Recurrence & Re-RT Treatment Details.

| Patient # | ECOG PS at time of Re-RT consult | Time from initial RT to PRDR (months) | Recurrence Site | Adjuvant or Definitive Re-RT | Indication for Re-RT | Area of Initial RT dose overlap | Re-RT Dose (cGy) & # of fractions | IMRT Technique | CTV Volume (cc) | Systemic Therapy |
| --- | --- | --- | --- | --- | --- | --- | --- | --- | --- | --- |
| 1 | 1 | 6 | Nodal | Adjuvant | ECE+ | Elective dose | 6000cGy in 30 fractions | Helical Tomotherapy | 30.5 | Concurrent & Adjuvant |
| 2 | 1 | 13 | Primary & Nodal | Adjuvant | Tumor Board Recommendation | High dose | 6000cGy in 30 fractions | Helical Tomotherapy | 79.8 | Concurrent |
| 3 | 2 | 17 | Nodal | Adjuvant | ECE+ | High dose | 6000cGy in 30 fractions | Helical Tomotherapy | 19.1 | Concurrent |
| 4 | 2 | 19 | Primary | Adjuvant | Tumor Board Recommendation | High dose | 6000cGy in 30 fractions | Helical Tomotherapy | 234.1 | None |
| 5 | 2 | 7 | Nodal | Adjuvant | Margin+ | High dose | 6000cGy in 30 fractions | Helical Tomotherapy | 46.45 | Concurrent & Adjuvant |
| 6 | 2 | 50 | Primary | Definitive | Refused Salvage Surgery | High dose | 6600cGy in 33 fractions | Helical Tomotherapy | 112.13 | Adjuvant |
| 7 | 2 | 10 | Primary & Nodal | Adjuvant | Tumor Board Recommendation | High dose | 6000cGy in 30 fractions | Helical Tomotherapy | 126.04 | None |
| 8 | 1 | 12 | Primary | Adjuvant | Margin+ | High dose | 6000cGy in 30 fractions | VMAT | 47.28 | None |
| 9 | 1 | 32 | Primary | Adjuvant | Tumor Board Recommendation | High dose | 6000cGy in 30 fractions | Helical Tomotherapy | 65.19 | None |

Re-RT = Re-irradiation ECOG PS = Eastern Cooperative Oncology Group Performance Status PRDR = Pulsed Reduced Dose Rate IMRT = Intensity Modulated Radiotherapy VMAT = Volume Modulated Arc Therapy CTV = Clinical Target Volume ECE = Extracapsular Extension

### Toxicity

Of acute toxicities attributed to PRDR Re-RT, the majority were grade 1 or 2. There were no acute grade 4 or 5 toxicities. One patient (11.1%) had acute grade 3 radiation dermatitis managed with silver sulfadiazine. In terms of late toxicities potentially attributable to PRDR Re-RT, the majority were again grade 1 or 2. Two patients (22.2%) had late grade 3 toxicities: one patient had esophageal stenosis requiring dilation, and the other patient had grade 3 xerostomia due to inability to close her mouth after salvage surgery (revision surgery is planned). There were no late grade 4 or 5 late toxicities. Toxicity events are detailed in Table 3.

**Table 3:** Peak Toxicity Rates for Patients Receiving PRDR Re-RT.

| **Toxicity** | **Number of toxicity events** |  |  |  |  |
| --- | --- | --- | --- | --- | --- |
|  | Grade 1 | Grade 2 | Grade 3 | Grade 4 | Grade 5 |
| **<u>Acute</u>** |  |  |  |  |  |
| Anorexia | 2 | 2 | 0 | 0 | 0 |
| Fatigue | 3 | 4 | 0 | 0 | 0 |
| Dermatitis | 4 | 1 | 1 | 0 | 0 |
| Nausea | 2 | 2 | 0 | 0 | 0 |
| Xerostomia | 3 | 1 | 0 | 0 | 0 |
| Dysphagia | 0 | 1 | 0 | 0 | 0 |
| Cervical Esophagitis | 1 | 1 | 0 | 0 | 0 |
| Oral Mucositis | 2 | 1 | 0 | 0 | 0 |
| Dysgeusia | 0 | 2 | 0 | 0 | 0 |
| **<u>Late</u>** |  |  |  |  |  |
| Anorexia | 0 | 2 | 0 | 0 | 0 |
| Fatigue | 3 | 1 | 0 | 0 | 0 |
| Dermatitis | 1 | 0 | 0 | 0 | 0 |
| Tinnitus | 1 | 0 | 0 | 0 | 0 |
| Xerostomia | 1 | 2 | 1 | 0 | 0 |
| Dysphagia | 1 | 1 | 0 | 0 | 0 |
| Trismus | 0 | 1 | 0 | 0 | 0 |
| Dysgeusia | 2 | 0 | 0 | 0 | 0 |
| Lymphedema | 1 | 2 | 0 | 0 | 0 |
| Fibrosis | 1 | 1 | 0 | 0 | 0 |
| Esophageal Stenosis | 0 | 0 | 1 | 0 | 0 |
| Soft tissue Necrosis | 0 | 1 | 0 | 0 | 0 |

PRDR = Pulsed Reduced Dose Rate
Re-RT = Re-irradiation

### Outcomes

The median follow-up after Re-RT was 7 months. In this time, 3 deaths were observed. One patient died due to unrelated pneumonia 1 month after treatment completion; this patient had not received chemotherapy and did not have risk of aspiration because he had a total laryngectomy. Two patients died due to in-field disease progression. The median OS from PRDR Re-RT was 7 months (range, 1-32 months). Overall, 3 patients had in-field progression: one at 6 months, one at 5 months and one at 3 months. Median PFS was 7 months (range, 1-32 months). Two patients are alive and disease-free 25+ months (25 and 32 months) from Re-RT completion.

## Discussion

To our knowledge, this single-institutional case series represents the largest group of patients with recurrent or second primary HNSCC treated with PRDR-RT for Re-RT. Data presented above demonstrated encouraging preliminary safety and efficacy outcomes for this challenging clinical situation.

Current treatment options for recurrent or second primary HNSCC after previous RT are limited. In terms of systemic therapy, the CheckMate 651 trial (which randomized patients to immunotherapy with nivolumab/ipilimumab versus combination chemotherapy with cetuximab) reported a median overall survival of 13.9 months.^12^ Salvage surgical resection (if possible) remains the standard of care.^10^ The benefit of adding Re-RT after surgery is unclear. The phase 3 GORTEC study by Janot and colleagues reported that adjuvant Re-RT after surgery improved DFS and LRC but not OS.^13^ There were significant rates of toxicity with acute grade 3-4 complications in 28% of cases and late grade 3-4 toxicities in 39%, which could have affected OS as deaths related to locoregional disease were actually decreased. However, this trial was conducted from 1999-2005 and employed older radiation techniques and fractionation schemes.

Modern series using contemporary RT techniques (standard dose rate IMRT, SBRT, proton therapy) report lower but still significant rates of toxicity. A pooled multi-institutional analysis of 412 HNSCC patients receiving Re-RT with standard dose rate IMRT/VMAT reported a rate of severe (grade ≥3) toxicity of 19%.^4^ Life-threatening (grade 4) acute toxicity occurred in 4.4% of patients; 1.2% of patients were thought to have grade 5 toxicity.^4^ A recent phase 1 trial examining the safety of SBRT for Re-RT reported 28% rate of acute grade 3 toxicity with 1 death scored as possibly related to treatment.^14^ Proton therapy is a means of reducing normal tissue dose; a recent series of proton Re-RT for recurrent HNSCC reported a late grade ≥3 toxicity rate of 32.6%; grade 4 and 5 toxicity occurred in 1.6% and 2.1% of patients, respectively.^15^

In terms of toxicity, our series of PRDR Re-RT compares favorably with these recently published Re-RT series. However, most of the patients in this study received Re-RT after salvage surgery which makes comparison to other re-irradiation series more difficult. Biologically, PRDR-RT is thought to target dividing neoplastic cells effectively while permitting intratherapy sublethal damage repair in previously irradiated normal tissue, which may decrease normal tissue toxicity while maintaining efficacy.^16^ This may provide an advantage for PRDR-RT compared to standard dose rate IMRT, SBRT, and proton therapy. A recently published trial examining PRDR-RT for Re-RT of recurrent NPC reported 1-year cancer specific survival rate of 80.6% as well as acute and late grade ≥3 toxicity rates of 22.2% and 19.4%, respectively.^17^ These rates were lower than historical controls for standard dose rate IMRT-based Re-RT, especially given the anatomic difficulties of treating NPC with Re-RT. These findings suggest the potential benefits of PRDR-RT in reducing Re-RT toxicity while maintaining efficacy. In addition, PRDR could be utilized in combination with SBRT or proton therapy to further improve toxicity outcomes, although this would be logistically challenging.

Limitations of our study include the small sample size and the biases involved with a retrospective review. However, this initial experience and toxicity evaluation suggests that Re-RT using PRDR-RT is a well-tolerated regimen and compares favorably to other treatment strategies for recurrent or second primary HNSCC. Our group is working on initiating a pilot prospective study to gather preliminary safety and efficacy data for PRDR-RT for Re-RT in this patient population.

## Conclusion

PRDR-RT for Re-RT is a feasible treatment strategy for patients with recurrent or second primary HNSCC. Initial findings from this retrospective review suggest reasonable survival outcomes and potentially improved toxicity; prospective data is needed to establish the safety and efficacy of this technique.

## Data Availability

Research data is not available at this time.

## Notes

**[Conflict of Interest Statement for All Authors]** The authors have no conflicts of interest to disclose.

### Competing Interest Statement

The authors have declared no competing interest.

### Funding Statement

This study did not receive any funding

### Author Declarations

Ethics committee/IRB of the University of Arkansas for Medical Sciences waived ethical approval for this work.

